# Identification of Immune complement function as a determinant of adverse SARS-CoV-2 infection outcome

**DOI:** 10.1101/2020.05.05.20092452

**Authors:** Vijendra Ramlall, Phyllis M. Thangaraj, Cem Meydan, Jonathan Foox, Daniel Butler, Ben May, Jessica K. De Freitas, Benjamin S. Glicksberg, Christopher E. Mason, Nicholas P. Tatonetti, Sagi D. Shapira

## Abstract

Understanding the pathophysiology of SARS-CoV-2 infection is critical for therapeutics and public health intervention strategies. Viral-host interactions can guide discovery of regulators of disease outcomes, and protein structure function analysis points to several immune pathways, including complement and coagulation, as targets of the coronavirus proteome. To determine if conditions associated with dysregulation of the complement or coagulation systems impact adverse clinical outcomes, we performed a retrospective observational study of 11,116 patients who presented with suspected SARS-CoV-2 infection. We found that history of macular degeneration (a proxy for complement activation disorders) and history of coagulation disorders (thrombocytopenia, thrombosis, and hemorrhage) are risk factors for morbidity and mortality in SARS-CoV-2 infected patients - effects that could not be explained by age, sex, or history of smoking. Further, transcriptional profiling of nasopharyngeal (NP) swabs from 650 control and SARS-CoV-2 infected patients demonstrated that in addition to innate Type-I interferon and IL-6 dependent inflammatory immune responses, infection results in robust engagement and activation of the complement and coagulation pathways. Finally, we conducted a candidate driven genetic association study of severe SARS-CoV-2 disease. Among the findings, our scan identified putative complement and coagulation associated loci including missense, eQTL and sQTL variants of critical regulators of the complement and coagulation cascades. In addition to providing evidence that complement function modulates SARS-CoV-2 infection outcome, the data point to putative transcriptional genetic markers of susceptibility. The results highlight the value of using a multi-modal analytical approach, combining molecular information from virus protein structure-function analysis with clinical informatics, transcriptomics, and genomics to reveal determinants and predictors of immunity, susceptibility, and clinical outcome associated with infection.

## Introduction

The SARS-CoV-2 pandemic has had profound economic, social, and public health impact with over 6.1 million confirmed cases and over 370,000 deaths across the globe. The infection causes respiratory illness with symptoms ranging from cough and fever to difficulty breathing. While highly variable age-dependent mortality rates have been widely reported, the comorbidities that drive this dependence are not fully understood. Further, with some notable exceptions^1-3^, molecular studies have largely focused on ACE-2, the receptor and determinant of cell entry and viral replication^3^. While ACE-2 expression is critical, viruses employ a wide range of molecular strategies to infect cells, avoid detection, and proliferate. In addition, viral replication and immune mediated pathology are the primary drivers of morbidity and mortality associated with SARS-CoV-2 infection^4,5^. Therefore, understanding how virus-host interactions manifest as SARS-CoV-2 risk factors will facilitate clinical management, choice of therapeutic interventions, and setting of appropriate social and public health measures.

Knowledge of the precise molecular interactions that control viral replicative cycles can delineate regulatory programs that mediate immune pathology associated with infection and provide valuable clues about disease determinants. For example, viruses, including SARS-CoV-2, deploy an array of genetically encoded strategies to co-opt host machinery. Among the strategies, viruses encode multifunctional proteins that harness or disrupt cellular functions, including nucleic acid metabolism and modulation of immune responses, through protein-protein interactions and molecular mimicry - structural similarity between viral and host proteins (for a full discussion please see accompanying paper). Recently, we employed protein structure modeling to systematically chart interactions across all human infecting viruses^6^ and in an accompanying paper, performed a virome-wide scan for molecular mimics. This analysis points to broad diversification of strategies deployed by human infecting viruses and identifies biological processes that underlie human disease. Of particular interest, we mapped over 140 cellular proteins that are mimicked by coronaviruses (CoV). Among these, we identified components of the complement and coagulation pathways as targets of structural mimicry across all CoV strains (see companion paper).

Through activation of one of three cascades, (i) the classical pathway triggered by an antibody-antigen complex, (ii) the alternative pathway triggered by binding to a host cell or pathogen surface, and (iii) the lectin pathway triggered by polysaccharides on microbial surfaces, the complement system is a critical regulator of host defense against pathogens including viruses^7^. When dysregulated by germline variants or acquired through age-related effects or excessive acute and chronic tissue damage, complement activation can contribute to pathologies mediated by inflammation^7-9^. Similarly, inflammation-induced coagulatory programs -- which themselves can be regulated by the complement system -- as well as crosstalk between pro-inflammatory cytokines and the coagulative and anticoagulant pathways play pivotal roles in controlling pathogenesis associated with infections. Therefore, while the age-related differences in susceptibility to SARS-CoV-2 are likely a consequence of multiple underlying variables, virally encoded structural mimics of complement and coagulation pathway components may contribute to CoV associated immune mediated pathology. Moreover, a corollary of these observations is that dysfunctions associated with complement and/or coagulation may impact clinical outcome of SARS-CoV-2 infection. For example, the companion study suggests that coagulation disorders, such as thrombocytopenia, thrombosis and hemorrhage, may represent risk factors for SARS-CoV-2 clinical outcome. Among complement-associated disorders, multiple genetic and experimental evidence (including animal models of disease, histological examination of affected tissue, and germline mutational analysis) point to dysregulation of the complement system as the major driver of both early-onset, and age-related macular degeneration (AMD)^8-11^. A hyperinflammatory phenotype mediated by complement leads to progressive immune-mediated deterioration of the central retina. While AMD, the leading cause of blindness in elderly individuals (affecting roughly 200 million people worldwide^11^), is likely the result of multiple pathological processes, dysregulation of complement activation has emerged as the most widely accepted cause of disease^9-12^.

To determine if conditions associated with dysregulation of the complement or coagulation systems impact adverse clinical outcomes associated with SARS-CoV-2 infection, we conducted a retrospective observational study of 11,116 patients at New York-Presbyterian/Columbia University Irving Medical Center. In agreement with previous reports^13^, survival analysis identified significant risk of mechanical respiration and mortality associated with age and sex, as well as history of hypertension, obesity, type 2 diabetes (T2D), and coronary artery disease (CAD). Moreover, we found that patients with history of macular degeneration (a proxy for complement activation disorders) and coagulation disorders (i.e. thrombocytopenia, thrombosis, and hemorrhage) were at significantly increased risk of adverse clinical outcomes (including mechanical respiration and death) following SARS-CoV-2 infection. Importantly, these effects could not be explained by either age or sex, nor did we find any evidence that history of smoking contributes to risk of adverse clinical outcomes associated with SARS-CoV-2 infection. Conversely, albeit in a small number of individuals, we observed that no patients with complement deficiency disorders required mechanical respiration or succumbed to their illness. In addition, transcriptional profiling of nasopharyngeal (NP) swabs from 650 control and SARS-CoV-2 infected patients demonstrates that in addition to innate Type-I interferon and IL-6 dependent inflammatory immune responses, infection results in robust engagement and activation of the complement and coagulation pathways.

Finally, a focused analysis of proximal and distal variants of complement and coagulation components using the April 2020 COVID data released by the UK Biobank revealed genetic markers associated with severe SARS-CoV-2 infection. Among our findings, we identified variants in CD55 (a negative regulator of complement activation^14^), CFH and C4BPA, which play central roles in complement activation and innate immunity. Importantly, analysis of the May 2020 COVID data released by the UK Biobank recapitulated these results and identified additional variants. For example, the scan revealed that variants in Alpha-2-macroglobulin (A2M), a protease inhibitor and cytokine transporter which participates in the formation of fibrin clots and regulates inflammatory cascades, were associated with adverse clinical outcome. In addition to providing evidence that complement function modulates SARS-CoV-2 infection, the data point to several putative genetic markers of susceptibility. The results highlight the value of using a multi-modal analytical approach, combining molecular information from virus protein structure-function analysis with clinical informatics, transcriptomics, and genomics to reveal determinants and predictors of immunity, susceptibility, and clinical outcome associated with infection.

## Results

### Comorbidity statistics and covariances in a retrospective observational clinical cohort

To explore if conditions associated with dysregulation of the complement or coagulation systems impact adverse clinical outcomes associated with SARS-CoV-2, we conducted a retrospective observational study of patients treated at New York-Presbyterian/Columbia University Irving Medical Center for suspected infection (Table 1). Electronic health records (EHR) were used to define sex, age, and smoking history status as well as histories of macular degeneration, coagulatory disorders (i.e. thrombocytopenia, thrombosis, and hemorrhage), hypertension, type 2 diabetes, coronary artery disease, and obesity (see Methods). As shown in Table 1, of the 11,116 patients that presented to the hospital between February 1, 2020 and April 25, 2020 with suspected SARS-CoV-2 infection, 6,398 tested positive for the virus. Among these, 88 were patients with a history of macular degeneration, four were patients with complement deficiency disorders, and 1,179 were patients with disorders associated with the coagulatory system. In addition, hypertension, coronary artery disease, diabetes, obesity, and annotated cough were represented by 1,922, 1,566, 847, 791, and 727 patients, respectively (Table 1). While CAD, hypertension, T2D, obesity, and coagulation disorders represent a group with the highest covariance, we find lower co-occurrence between these conditions and macular degeneration in both SARS-CoV-2 positive and negative individuals (Figure S1). In addition to these medical histories, smoking status, past or present, was noted for 5,079 patients (of 1,359 smokers included in the study, 723 were SARS-CoV-2 positive). Finally, of patients who were put on mechanical ventilation, we observed a 35% mortality rate, and 31% of deceased patients had been on mechanical respiration.

**Table 1.**
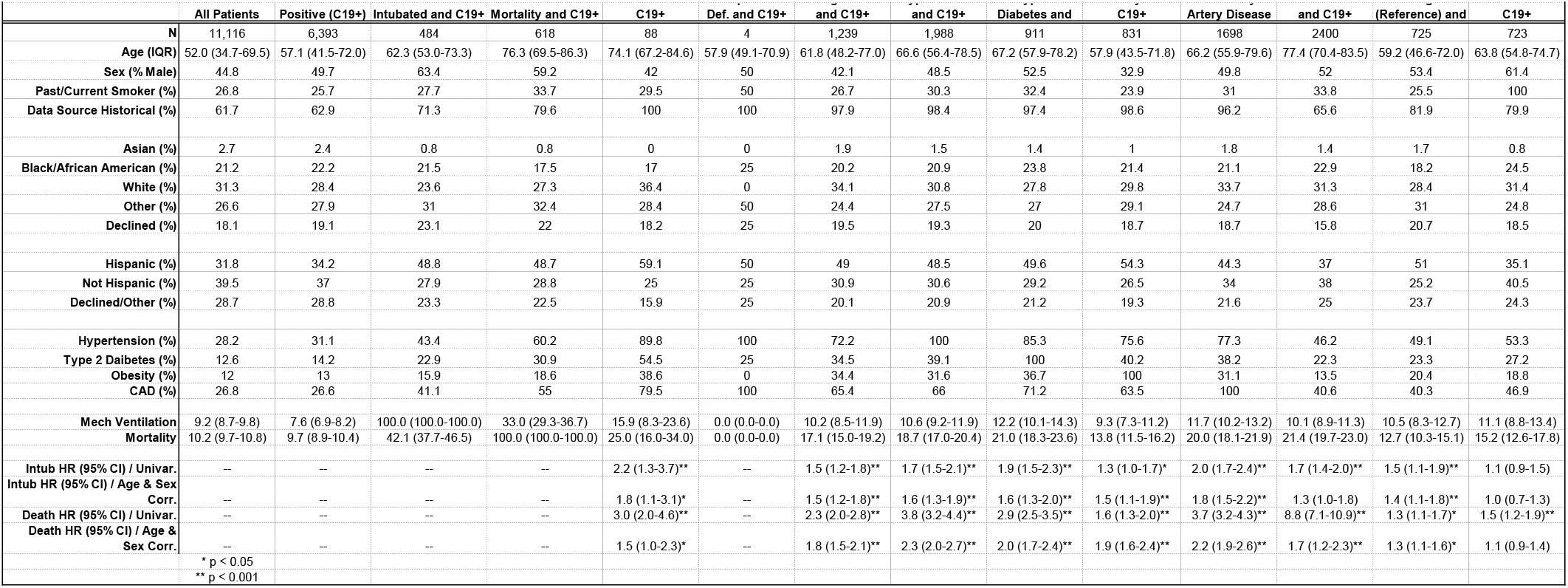
Cohort demographics and outcome associations in patients suspected of SARS-CoV-2 infections

### Macular degeneration and coagulation disorders are associated with SARS-CoV-2 outcomes

We estimated the univariate and age- and sex-corrected risk associated with baseline clinical history of previously reported SARS-CoV-2 risk factors (including hypertension, obesity, type 2 diabetes, and coronary artery disease) as well as coagulation and complement disorders using survival analysis and Cox proportional hazards regression modeling. As shown in Figure 1 and Table 1, we identified significant risk of mechanical respiration and mortality associated with age and sex, as well as history of hypertension, obesity, and type 2 diabetes (T2D), coronary artery disease (CAD). Notably, we did not find evidence that smoking status (past or present) is a significant risk factor for either mechanical respiration or mortality. We found that those with a history of macular degeneration (a proxy for complement activation disorders) and coagulation disorders (thrombocytopenia, thrombosis, and hemorrhage) were at significantly increased risk of adverse clinical outcomes (including mechanical respiration and death) following SARS-CoV-2 infection (Figure 1, Table 1). Specifically, we observed a mechanical respiration rate of 15.9% (95% CI: 8.3-23.6; HR: 2.2, *P*value = 0.0046) and a mortality rate of 25% (95% CI: 16.0-34.0; HR 3.0, *P*value = 4.4×10^-7^) among patients with a history of macular degeneration, and rates of 9.4% (95% CI: 7.7-11.1; HR 1.5, *P*value = 9.6×10^-5^) and 14.7% (95% CI: 12.7-16.7; HR: 2.3, *P*value = 1.8 ×10^-23^) for mechanical respiration and mortality, respectively, among patients with coagulation disorders (Table 1). Moreover, as shown in Figure 1b, patients with a history of macular degeneration appear to succumb to disease more rapidly than others. Critically, the contribution of age and sex was not sufficient to explain the increased risks associated with history of macular degeneration (Age/Sex-Corrected mechanical respiration HR=1.8 95% CI: 1.1-3.2, *P*value = 0.024; Age/Sex-Corrected mortality HR=1.7 95% CI: 1.1-2.5, *P*value = 0.022) or coagulation disorders (Age/Sex-Corrected mechanical respiration HR= 1.5. 95% CI: 1.2-1.8, *P*value = 2.4×10^-4^; Age/Sex-Corrected mortality HR=1.8 95% CI: 1.5-2.1, *P*value = 3.4×10^-12^). Conversely, albeit in a small number of individuals, we observed that among patients with complement deficiency disorders, who are normally at increased risk of complications associated with infections, none required mechanical respiration or succumbed to their illness (Table 1, Figure 1a and 1b). Importantly, while the correlation between macular degeneration or coagulopathies and established covariates included in this study is low (as shown in Supplemental Figure S1 and Supplemental Table S1, Tanimoto coefficients between 0.038 and 0.050 and 0.25 and 0.38, respectively), further study, perhaps with larger patient cohorts, will be necessary to rule out comorbidities that may be associated with macular degeneration and coagulopathies. Together, these data suggest that hyper-active complement and coagulative states predispose individuals to adverse outcomes associated with SARS-CoV-2 infection, and that deficiencies in complement components may be protective. Importantly, given the low incidence rate of deficiencies in either complement or coagulation pathways, further analysis with larger clinical cohorts is warranted.

**Figure 1.**
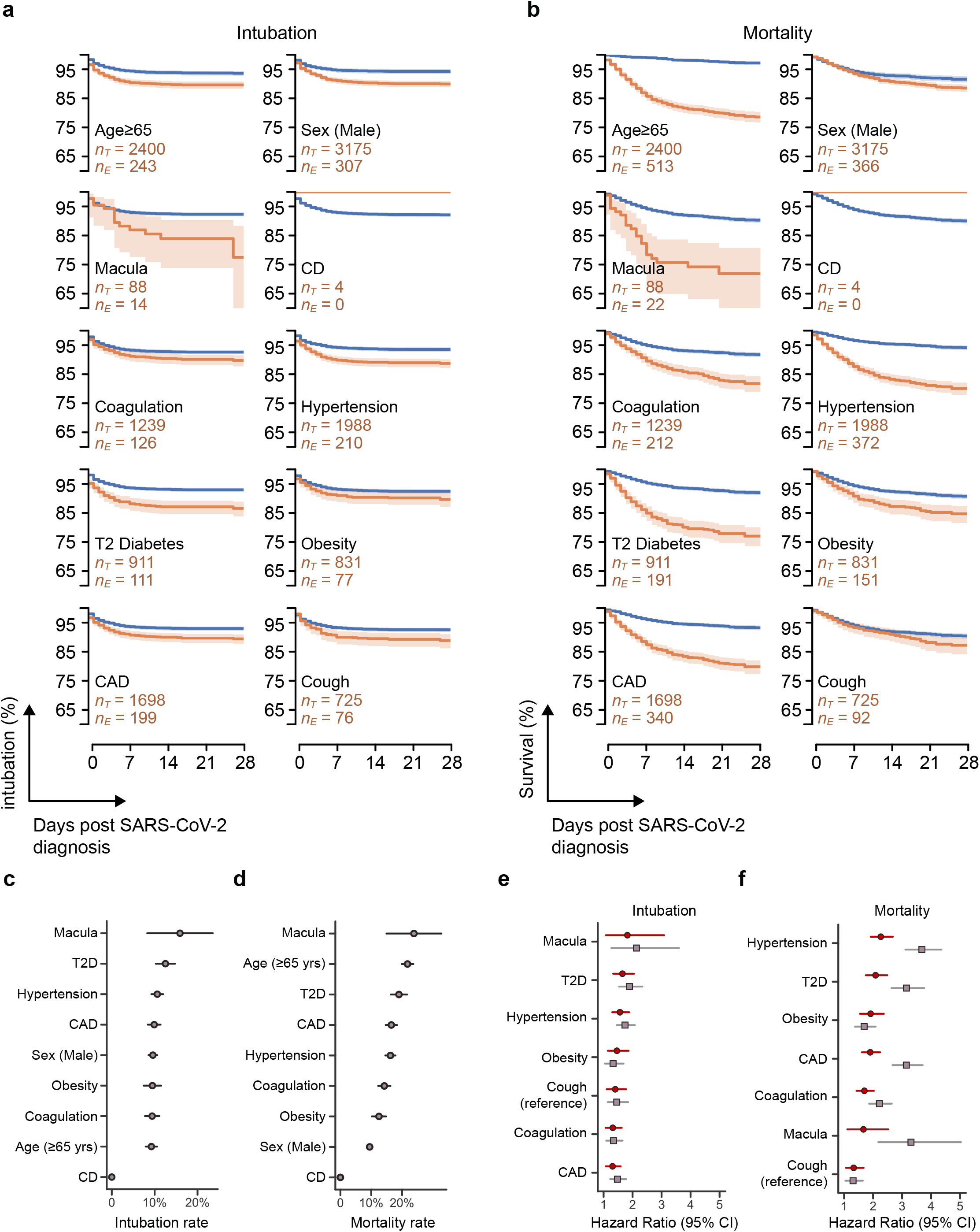
| History of macular degeneration and coagulation disorders are associated with adverse outcomes after confirmed SARS-CoV-2 infection. **a**, Kaplan-Meier curves for 10 binary conditions: age over 65, male sex, macular degeneration (Macula), complement deficiency disorders (CD), coagulation, hypertension, type 2 diabetes (T2DM), obesity, coronary artery disease (CAD), and cough. The survival for the patients with the named condition are shown in orange. The shaded region indicates the 95% confidence interval. The blue survival line is for patients without the named condition. Note that none of the four patients with CD required mechanical ventilation. **b**, Kaplan-Meier curves for the same 10 conditions as in (**a**). All four patients with CD survived (not statistically significant). **c**, Intubation rates across the binary conditions. Mortality (N=88) was highest in patients with a history of macular degeneration, followed by Type 2 Diabetes and Hypertension. **d**, Mortality rates across the binary conditions. Patients with a history of macular degeneration saw the highest mortality rates, followed by Age ≥ 65 and Type 2 Diabetes. **e**, Hazard ratios, estimated using a Cox proportional hazards model, for risk if intubation (as a validated proxy for requiring mechanical respiration). **f**, Similarly, hazard ratios for mortality, estimated using a Cox proportional hazards model. Hazard ratios and statistical significances are shown in Table 1.

### SARS-CoV-2 infection induces robust transcriptional regulation of complement and coagulation components

Transcriptional responses of human NP epithelial cells during viral infection can provide critical information about underlying immune programs. We leveraged whole genome RNA sequencing (RNA-seq) profiles to identify differentially regulated genes and pathways in 650 NP swabs from control and SARS-CoV-2 infected patients who presented to Weill-Cornell Medical Center. As shown in Figure 2a, gene set enrichment analysis (GSEA) of HALLMARK gene sets found that SARS-CoV-2 infection (as defined by presence of SARS-CoV-2 RNA and stratified into ‘positive’, ‘low’, ‘medium’ or ‘high’ based on viral load; see Methods) induces genes related to pathways with known immune modulatory functions, including ‘inflammatory_response’, ‘interferon_alpha_response’, and ‘IL6_JAK_STAT3_signaling (FDR corrected *P*value < 0.001; Figure 2a). Moreover, we found that among the most enriched gene sets, SARS-CoV-2 infection induces robust activation of the complement cascade (FDR corrected *P*value < 0.001), with increasing enrichment and significance with viral load (FDR corrected *P*value < 0.0001). We extended the analysis to include all complement and coagulation associated gene sets in MsigDB and identified ‘KEGG_Complement_and_Coagulation_Cascades’, ‘GO_Coagulation’, as well as ‘Reactome_initial_triggering_of_complement’ to be enriched in expression profiles of SARS-CoV-2 infected samples (*Q*value < 0.05; representative GSEA profiles are shown in Figure 2b and a full list of enriched pathways and gene sets can be found at https://masonlab.shinyapps.io/CovidGenes/). As highlighted in Figure 2c-e, the pathway-level transcriptional regulation induced by SARS-CoV-2 identified by GSEA is also observed at the individual gene level for upregulated and downregulated regulated transcripts as well as those that are particularly upregulated in the context of high viral load (Figure 2d, e, f, respectively). Taken together, the data demonstrate that in addition to immune factors like Type I interferons and dysregulation of IL6-dependent inflammatory responses which has been linked to poor clinical outcome^13^, transcriptional control of complement and coagulation cascades is a feature of SARS-CoV-2 infection.

**Figure 2.**
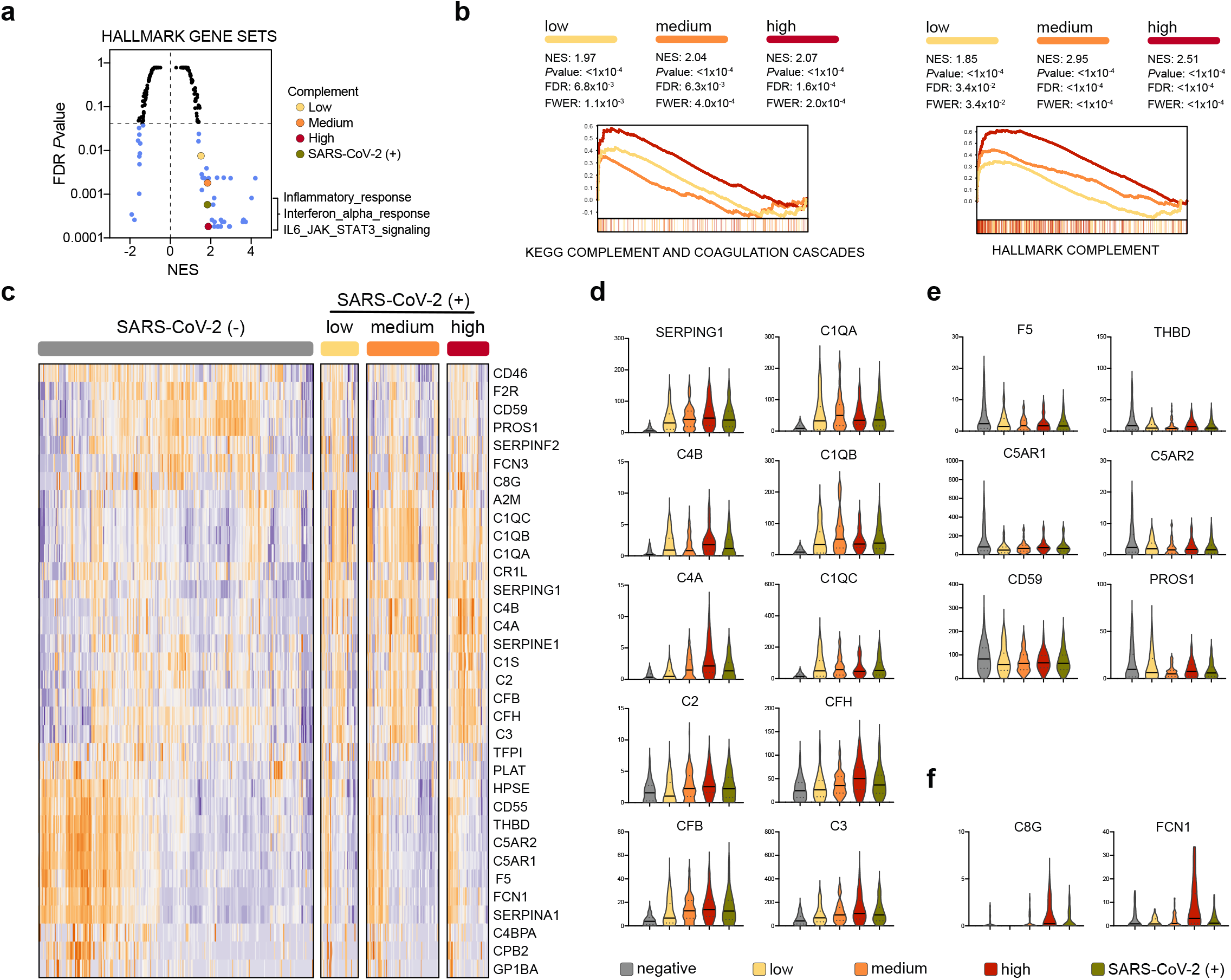
| SARS-CoV-2 infection engages robust transcriptional regulation of complement and coagulation cascades. **a**, GSEA of HALLMARK gene sets was applied to RNA-seq profiles of NP swabs from 650 control and SARS-CoV-2 infected patients stratified by SARS-CoV-2 positive (green) or low (yellow), medium (orange), high (red) viral load (significantly enriched gene sets highlighted in blue; **b**, Leading edge enrichment plots from GSEA analysis of MsigDB-wide gene sets are shown for HALLMARK_Complement and KEGG_Complement_and_Coagulation_Cascade gene sets with SARS-CoV-2 stratification indicated by color. **c**, Hierarchical clustering of Z-score normalized mRNA profiles of complement and coagulation components that undergo significant (FDR corrected *P*value < 0.01) transcriptional regulation in response to SARS-CoV-2 infection (cold and hot color scale reflects down, or up regulated expression, respectively). **d-f**, Violin plots (transcripts per million; TPM shown on *y*-axis) of highlighted differentially regulated genes are shown for upregulated (**d**), downregulated (**e**), or particularly upregulated in the context of high viral load (**f**). Normalized enrichment scores (NES) and FDR-corrected *P*values are shown.

### Genetic variation in complement and coagulation pathway components is associated with adverse SARS-CoV-2 infection outcome

The data highlighted above provide evidence that complement and coagulation disorders play a role in SARS-CoV-2 infection outcome and that infection with this virus induces robust transcriptional regulation of complement and coagulation pathway components. Moreover, dysfunction of complement or coagulation cascades can be the result of either acquired dysregulation, genetically encoded variants, or both. However, any genetic factors that may underlie the clinical trends we observed remain hidden due to the retrospective nature of the study and the lack of available genetic data on these patients. On the other hand, the UK Biobank, a prospective cohort study with deep genetic, physical, and health data collected on ~500,000 individuals across the United Kingdom^15,16^, recently released SARS-CoV-2 infection and outcome statuses for 1,474 patients, allowing for genetic and epidemiological associations to be assessed. The release in April 2020 included 669 patients who tested positive for the virus, 572 of whom required hospitalization.

We conducted a candidate driven study to evaluate if genetic variation in components of complement or coagulation pathways are associated with poor SARS-CoV-2 clinical outcome. Briefly, we focused our analysis on 337,147 (181,032 female) subjects of White British descent, excluding 3rd degree and above relatedness and without aneuploidy^15^. Applying these restrictions to the April-2020 cohort resulted in 910 patients with suspected infection (388 positive, 332 positive and hospitalized; see *Methods)*. As detailed Supplemental Table S2, of the 805,426 genetic variants profiled in the UK Biobank, 2,888 are within a 60Kb window around 102 genes with known roles in regulating complement or coagulation cascades (results that follow are robust to varying window size between 40Kb-80Kb; see *Methods*, Figure 3a-b). We focused our analysis on single-nucleotide polymorphisms (SNP) with minor allele frequency (MAF) above 1% and, as shown in Figure 3 and Supplemental Figure S2a-f, used an empirical permutation analysis to set the study-wide significance alpha (α) thresholds for each analysis described below (see *Methods)*. As highlighted in Figure 3c and further detailed in Supplemental Table S2, we identified 11 loci representing 7 genes with study-wide significance (α = 0.001) in the April-2020 cohort. Among these, and proximal to coagulation factor III (F3), is variant rs72729504 which we find to be associated with increased risk of adverse clinical outcome associated with SARS-CoV-2 infection (OR: 1.93). Fibrin fragment D-dimer, one of several peptides produced when cross-linked fibrin is degraded by plasmin, is the most widely used clinical marker of activated blood coagulation. Among the genetic loci that influence D-dimer levels, GWAS studies have identified mutations in F3 as having the strongest association^17^. Importantly, increased D-dimer levels were recently reported to correlate with poor clinical outcome in SARS-CoV-2 infected patients^13^. So, while the functional role of rs72729504 remains to be elucidated, our observations suggest that this locus may represent a genetic marker of SARS-CoV-2 susceptibility and outcomes.

**Figure 3.**
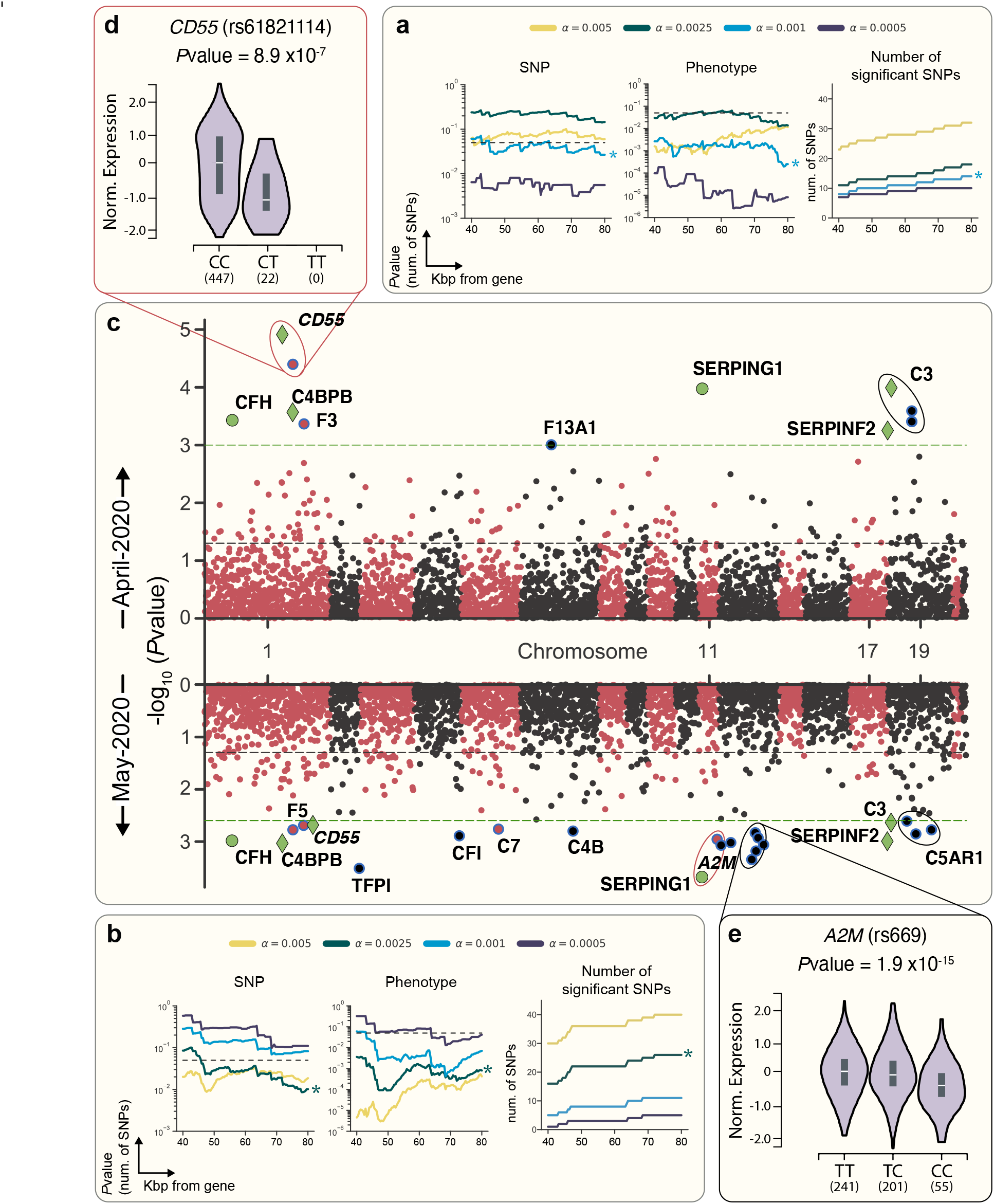
| Targeted genetic association study identifies SNPs in complement and coagulation pathway components associated with clinical outcome of SARS-CoV-2 infection. **a-b**, *P*values from a Negative Binomial distribution fit to permutation of SNPs sampled (left) and case:control phenotypes (center) generated under the null hypothesis are shown for the April-2020 (**a**) or May-2020 (**b**) cohort (a and distance pairs as indicated; for more information see *Methods*). Also shown are the number of hits that pass the corresponding alpha study-wide significance threshold by distance (right) for April-2020 (**a**) or May-2020 (**b**) cohorts. **c**, Manhattan plots of 2,888 variants within 60kb of complement and coagulation pathway genes for analyses using the April-2020 cohort (top) and May-2020 cohort (bottom). Study-wide significance threshold shown as dashed green lines, nominal significance threshold shown as black dashed line, and SNPs color alternates by chromosome. Significant SNPs are shown as colored markers and annotated with the nearest gene by base-pair distance. SNPs shown in green are study-wide significant in both April-2020 and May-2020. SNPs shown as diamonds are also study-wide significant in haplotype-based analysis (see *Methods*). eQTLs are further highlighted in (**d**) and (**e**). **d**, eQTL relationship for rs61821114 and *CD55* in thyroid^19^. The T allele of rs61821114 is associated with significantly lower expression of *CD55*. **e**, eQTL relationship for rs669 and *A2M^19^*. The C allele of rs669 is associated with significant lower expression of *A2M* in 17 tissues, including the esophageal mucosa (shown) and lung.

In addition to the SNP highlighted above, we identified 4 variants (rs45574833, rs61821114, rs61821041, and rs12064775) previously reported as risk alleles for AMD in the UKBB dataset^18^. Moreover, we find that each of these variants predisposes carriers to adverse clinical outcome (i.e. hospitalization) following SARS-CoV-2 infection (OR: 2.13-2.65). A fifth variant, rs2230199, which maps to complement C3, was shown to be linked to AMD in an independent GWAS, however, this variant has not been associated with increased AMD risk in the UK population. The three SNPs that map to C3 each appear to confer some protection associated with SARS-CoV-2 infection (OR: 0.66-0.68). In addition, two of the identified variants (rs61821114 and rs61821041) map to expression quantitative trait loci (eQTL) associated with Complement Decay-Accelerating Factor (CD55)^19^. This protein negatively regulates complement activation by accelerating the decay of complement proteins, thereby disrupting the cascade and preventing immune-mediated damage^7^. As reported by GTex Consortium data^19^ and highlighted in Figure 3d, these eQTLs result in decreased expression of CD55, thereby relieving the restraining function of this protein. In agreement with the functional role of CD55, we observe that these variants are associated with increased risk of adverse clinical outcome associated with SARS-CoV-2 infection (OR: 2.34-2.4).

Genetic association studies performed on relatively small cohorts can be prone to false positives. While permutation analyses to empirically determine statistical significance thresholds were implemented as described in *Methods*, we also repeated the analysis using updated UKBB data released in May, 2020 which included 3,002 patients with suspected infection. Of the 1,073 that tested positive in the updated cohort, 818 required hospitalization (651 and 500 respectively, after ancestry and relatedness filtering, see Methods). Importantly, analysis of the May-2020 COVID data recapitulated 6 of 11 April-2020 findings and identified 16 additional loci with study-wide significance (α = 0.0025, Supplemental Table S2, Figure 3c). Among these, the scan revealed 5 variants proximal to Alpha-2-macroglobulin (A2M), a protease inhibitor and cytokine transporter which participates in the formation of fibrin clots and regulates inflammatory cascades^20^. Of these, 3 (rs10842898, rs669, and rs4883215) are eQTLs associated with significant downregulation of A2M (and concomitant upregulation of A2M-AS1, the antisense RNA of A2M; data available on gtextportal.org) in multiple tissues including mucosa of the esophagus (Pvalue = 1.9×10^-15^) as highlighted in Figure 3e. In addition to A2M, rs10842898 and rs669 are splicing quantitative trait loci (sQTLs) for Mannose-6-Phosphate Receptor (M6PR) a P-type lectin that regulates lysosomal cargo loading and participates in cellular responses to wound healing, cell growth and viral infection^21^ - suggesting that the SNPs identified may contribute to complex regulation of transcripts with immunological and antiviral roles.

As detailed in Supplemental Table S2, 936 of the variants that were part of the study are within haplotype blocks of analyzed genes (see *Methods*). Analysis focused on SNPs in complement and coagulation haplotype blocks (based on linkage disequilibrium; LD, See Methods) resulted in 16 study-wide significant SNPs (α = 0.01, Figure S3) using the April-2020 cohort, of which 8 repeated at study-wide significance (α = 0.0075, Figure S3) using the May-2020 dataset. These include rs45574833, a variant highlighted above that results in a missense mutation in C4BPA, a protein that controls activation of the classical complement pathway by mediating hydrolysis of complement factor C4b and degradation of the C3 convertase^22^ (see Supplemental Table S2). In addition, the haplotype-based analysis identified a link between rs731034 (an eQTL in Collectin Subfamily Member 11; COLEC11) and poor clinical outcome in both April-2020 (OR: 1.27) and May-2020 (OR: 1.33) cohorts. COLEC11, a member of the collectin family of C-type lectins, plays an important role in the innate immune system by binding to carbohydrate antigens (with a preference for fucose and mannose) on microorganisms including viruses, facilitating their recognition and removal. This eQTL variant results in significant upregulation of COLEC11 across multiple tissues including lung (*P*value = 1×10^-11^) and suggests that sugar moieties on viral proteins may serve as antigenic targets of immunological responses to SARS-CoV-2 infection. Though experimental validation and functional interrogation of the variants we have identified is required to elucidate their precise pathophysiology, taken together, our observations point to genetic variation in complement and coagulation components as a contributing factor in SARS-CoV-2 mediated disease.

## Discussion

Zoonotic infections like the SARS-CoV-2 pandemic pose tremendous risk to public health and socioeconomic factors on a global scale. While the innate and adaptive arms of the immune system are exquisitely equipped to deal with noxious agents including viruses, interactions between emerging pathogens and their human hosts can manifest in unpredictable ways. In the case of SARS-CoV-2 infection a combination of viral replication and immune mediated pathology are the primary drivers of morbidity and mortality. While recent analysis of coronavirus patients in China, suggests that high serum levels of interleukin-6 (IL-6), a proinflammatory cytokine, is associated with poor prognosis^13^ (and as shown in Figure 2, found to be transcriptionally regulated in SARS-CoV-2 patients) further delineation of the regulatory programs that mediate immune pathology associated with SARS-CoV-2 infection is necessary. As illustrated in the accompanying paper and by the results presented herein, knowledge of molecular interactions between virus and host can refine hypothesis-driven discovery of disease determinants.

Our scan for virus-encoded structural mimics across Earth’s virome pointed to molecular mimicry as a pervasive strategy employed by viruses and indicated that the protein structure space used by a given virus is dictated by the host proteome (see accompanying paper). Moreover, observations about how coronaviruses exploit this strategy provided clues about the cellular processes driving pathogenesis. Together with knowledge that CoV infections, including the SARS-CoV outbreak in 2002-2003 and the current SARS-CoV-2 outbreak^13^, result in hyper-coagulative phenotypes^23^, our protein structure-function analysis led us to hypothesize that conditions associated with complement or coagulatory dysfunction may influence outcomes of SARS-CoV-2 infections. Of these, among the most common are AMD (which is associated with hyper-activation of the complement pathway) and hyper-coagulative disorders. Their relatively high incidence rates together with SARS-CoV-2 prevalence in and around New York City made them reasonable candidates for a retrospective clinical study.

As presented above, in addition to rediscovering previously identified risk factors including age, sex, hypertension, and CAD we found that history of macular degeneration or coagulatory dysfunctions predispose patients to poor clinical outcomes (including increased risk of mechanical ventilation and death) following SARS-CoV-2 infection. Complement deficiencies on the other hand, appear to be protective. Their low incidence rates, however, make for a small sample size and invite further investigation. Moreover, retrospective studies of observational data have notable limitations in their data completeness, selection biases, and methods of data capture. As a result, claims on causality cannot be made - nor can we definitively rule out other clinical factors as possible drivers. Nevertheless, in an orthogonal analysis of 650 transcriptional profiles of NP swabs, we demonstrate that in addition to immune factors like Type I interferons and dysregulation of IL-6-dependent inflammatory responses, SARS-CoV-2 infection results in engagement and robust activation of complement and coagulation cascades. Dysregulation of complement and coagulation pathways leading to pathology resulting from viral infection is not without precedent. Indeed, it has been associated with Dengue virus infection where immune mediated pathology and dysregulation of complement is correlated with disease severity and mirrors that of acute SARS-CoV-2 disease^24^. Moreover, though different from the variants identified in this study, polymorphisms and haplotypes in CFH have been associated with severity of Dengue infection^25^, suggesting that complement and coagulatory disfunctions may represent risk factors for a broader range of pathogens.

Finally, since complement and coagulative dysfunctions can have both acquired and congenital etiologies, we implemented a focused, candidate-driven analysis of UK Biobank data to evaluate linkage between severe SARS-CoV-2 disease and genetic variation associated with complement and coagulation pathways. Our analysis identified putative complement and coagulation associated loci including missense, eQTL and sQTL variants of critical regulators of the complement and coagulation cascades. Though interpretation of these findings may be limited by sample size, site-specific biases in clinical care decisions, ancestral homogeneity and population stratification in the biobank data, and socioeconomic status of affected populations, to our knowledge, this is the first study to identify complement and coagulation functions as underlying risk-factors of SARS-CoV-2 disease outcome. In addition, given an existing menu of immune-modulatory therapies that target complement and coagulation pathways, the discovery provides a rationale to investigate these options for the treatment of SARS-CoV-2 associated pathology. Indeed, the therapeutic potential of complement modulation was recently introduced and further shown to be of significant benefit in a cohort of SARS-CoV-2 patients^26,27^.

Our study highlights the value of combining molecular information from virus protein structure-function analysis with orthogonal clinical data analysis to reveal determinants and/or predictors of immunity, susceptibility, and clinical outcome associated with infection. Such a framework can help refine large-scale genomics efforts and help power genomics studies based on informed biological and clinical conjectures. While identification of CoV encoded structural mimics guided the retrospective clinical studies, a molecular and functional link between those observations and our discovery of complement and coagulation functions as risk factors for SARS-CoV-2 pathogenesis remains to be elucidated. Nevertheless, the findings advance our understanding of how SARS-CoV-2 infection leads to disease and can help explain variability in clinical outcomes. Among the implications, the data warrant heightened public health awareness for individuals most vulnerable to developing adverse SARS-CoV-2 mediated pathology.

## Data Availability

Our data from NYP/CUIMC are protected by HIPAA and cannot be released. UK Biobank data are available through their application process. In addition, we are updating online summary data as additional patient data become available.

## Acknowledgements

This work was funded by NIH grants 5R01GM109018 and 5U54CA209997 to SS, R35GM131905 to NPT, F30HL140946 to PT, and equipment grants S10OD012351 and S10OD021764 to the Columbia University Department of Systems Biology. CEM would like to thank the Scientific Computing Unit (SCU), XSEDE Supercomputing Resources, the Starr Cancer Consortium (I13-0052), and funding from the WorldQuant Foundation, The Pershing Square Sohn Cancer Research Alliance, NASA (NNX14AH50G, NNX17AB26G), the National Institutes of Health (R21AI129851, R01MH117406, R01AI151059

## Declaration of interests

The authors declare no competing interests

**Figure S1.**
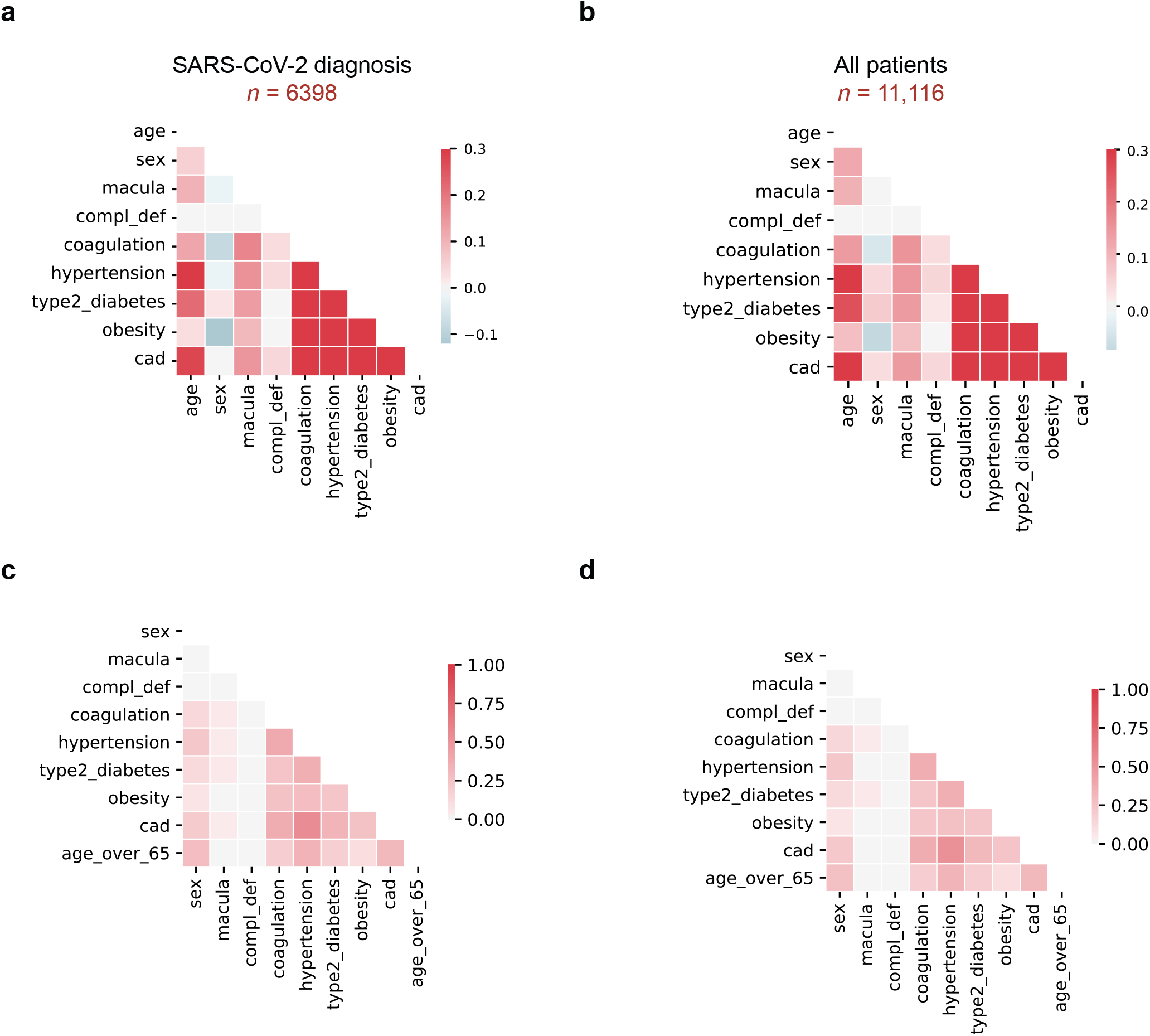
| Covariate correlations in EHR clinical data. **a**, Spearman correlation between modeled covariates in patients were diagnosed or tested positive for SARS-CoV-2: age, sex, macular degeneration (macula), complement deficiency disorders (CD), coagulation disorders (coagulation), hypertension, Type 2 Diabetes, obesity, and coronary artery disease (CAD). **b**, Spearman correlations, as in (**a**), for all patients (includes patients who tested negative for SARS-CoV-2). **c**, Tanimoto coefficients as in **(a)**, for patients who tested positive for SARS-CoV-2 infection. Age was binarized as “Age over 65” to compute the score. **d**, Tanimoto coefficients as in **(c)** for all patients.

**Figure S2.**
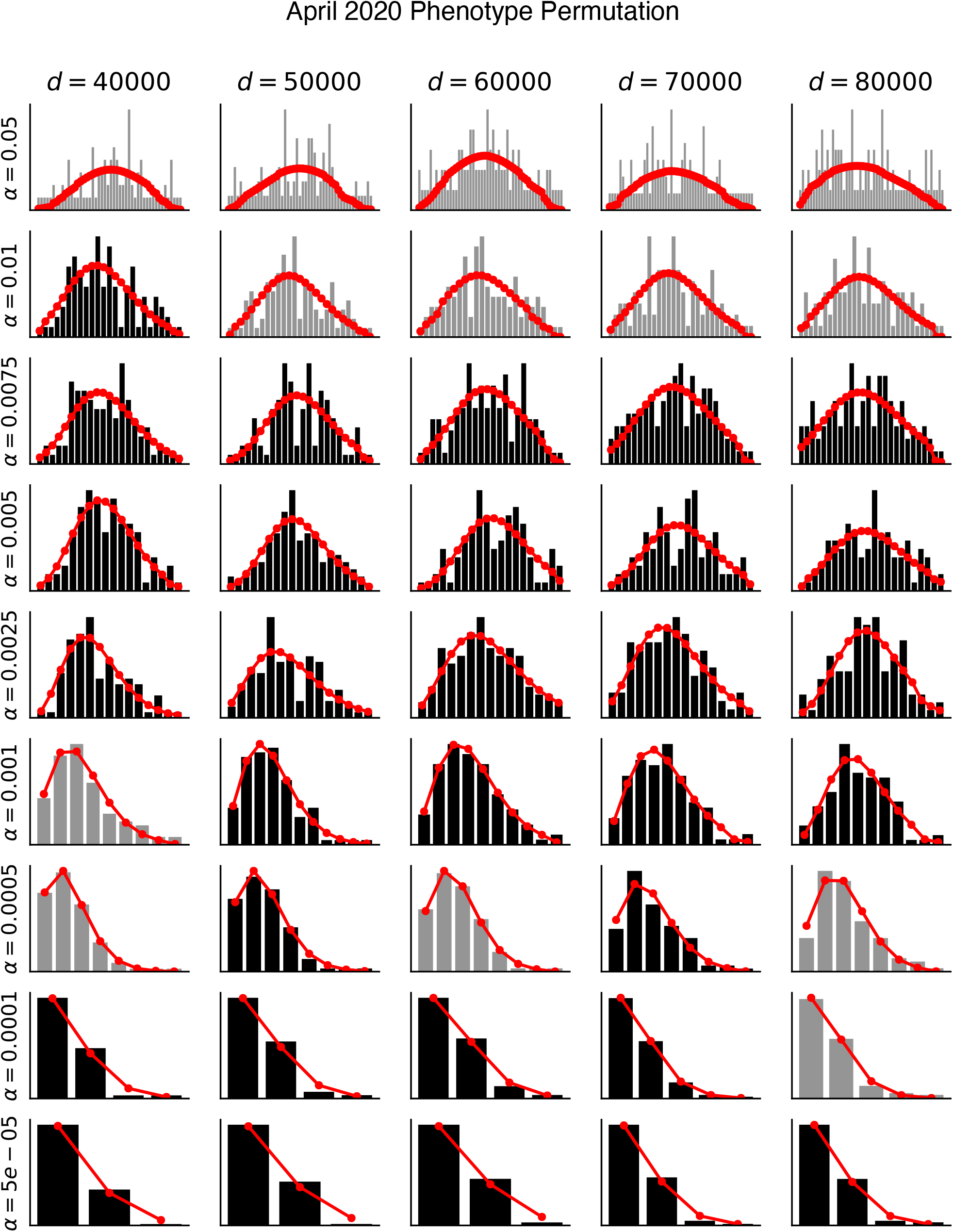
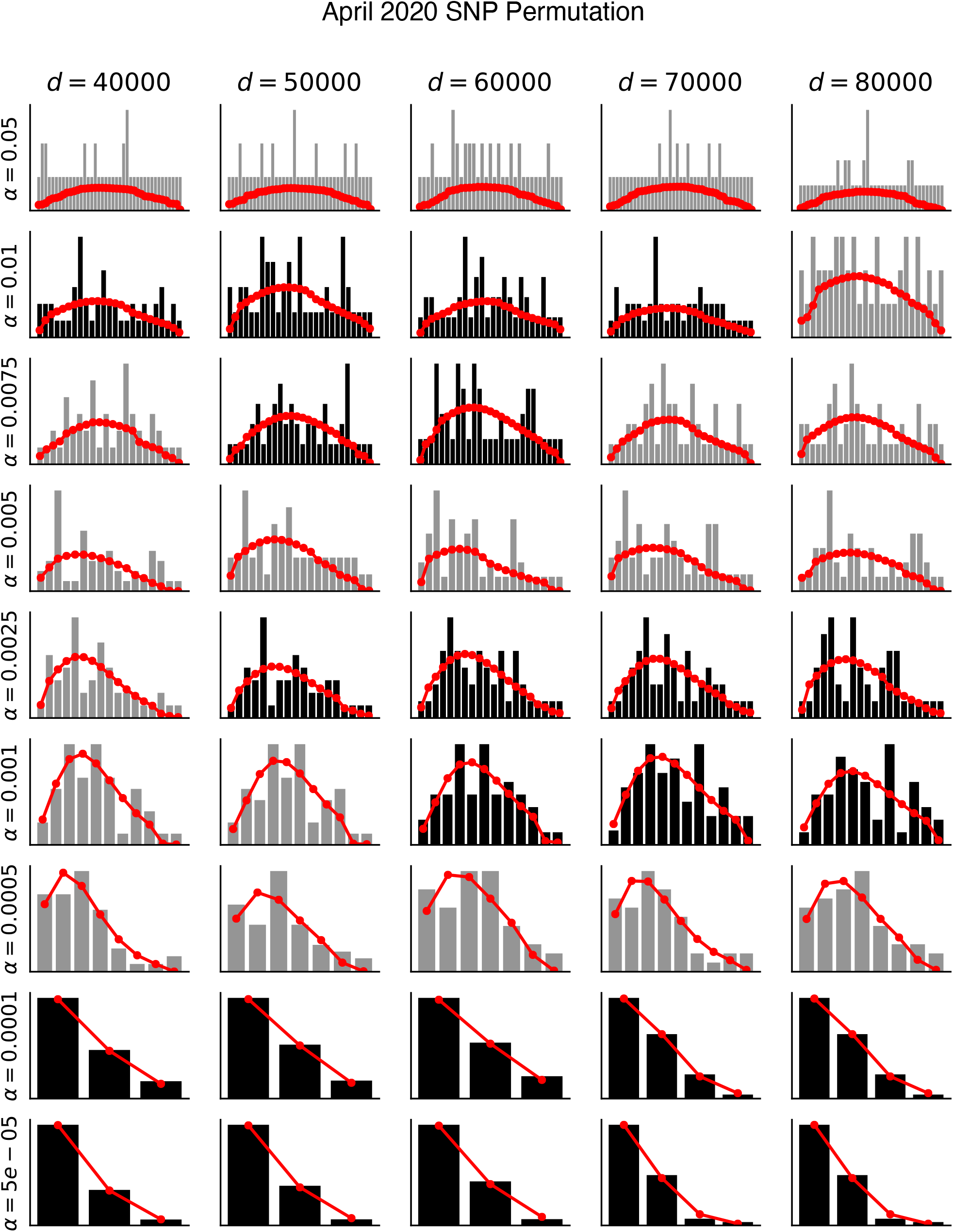
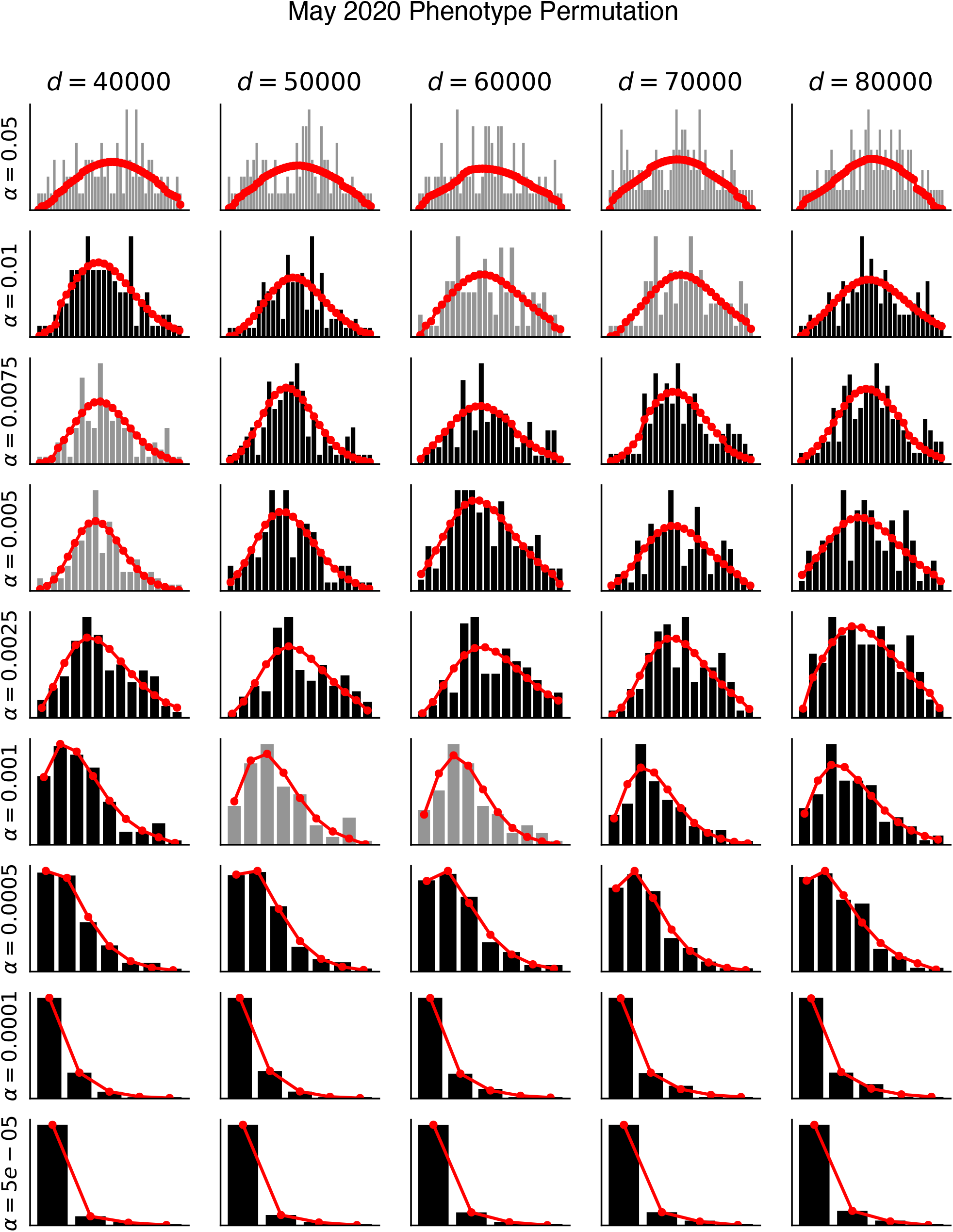
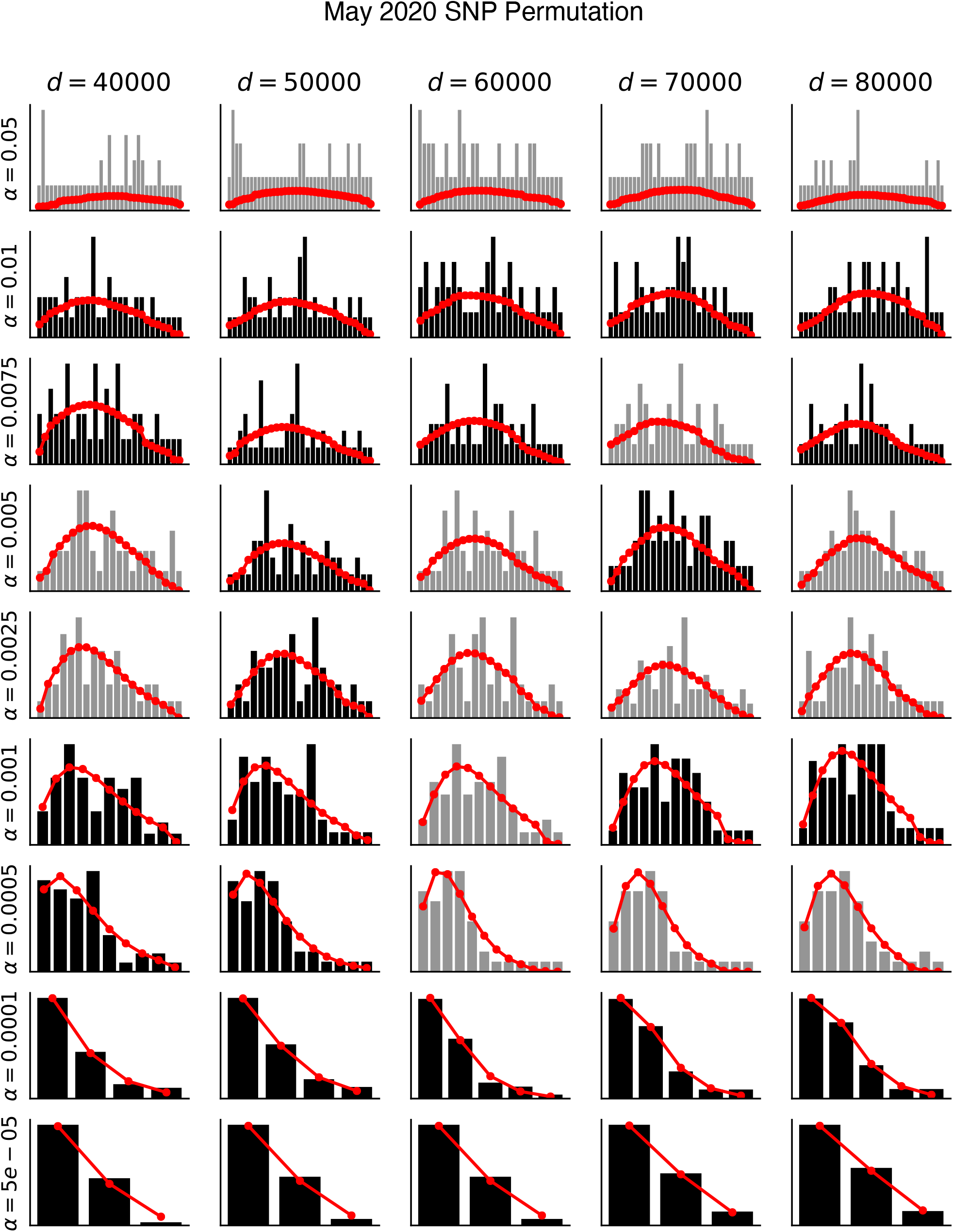
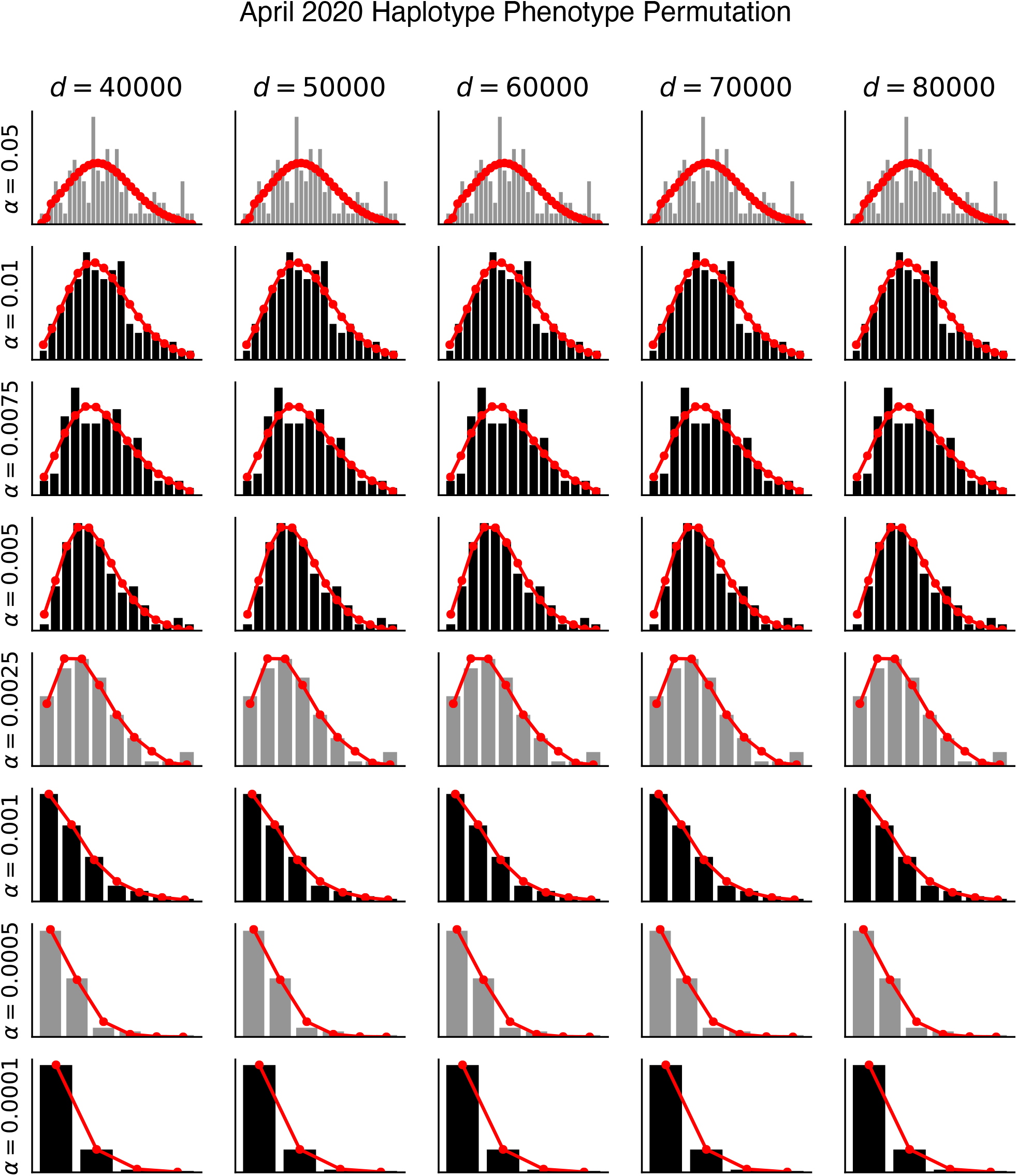
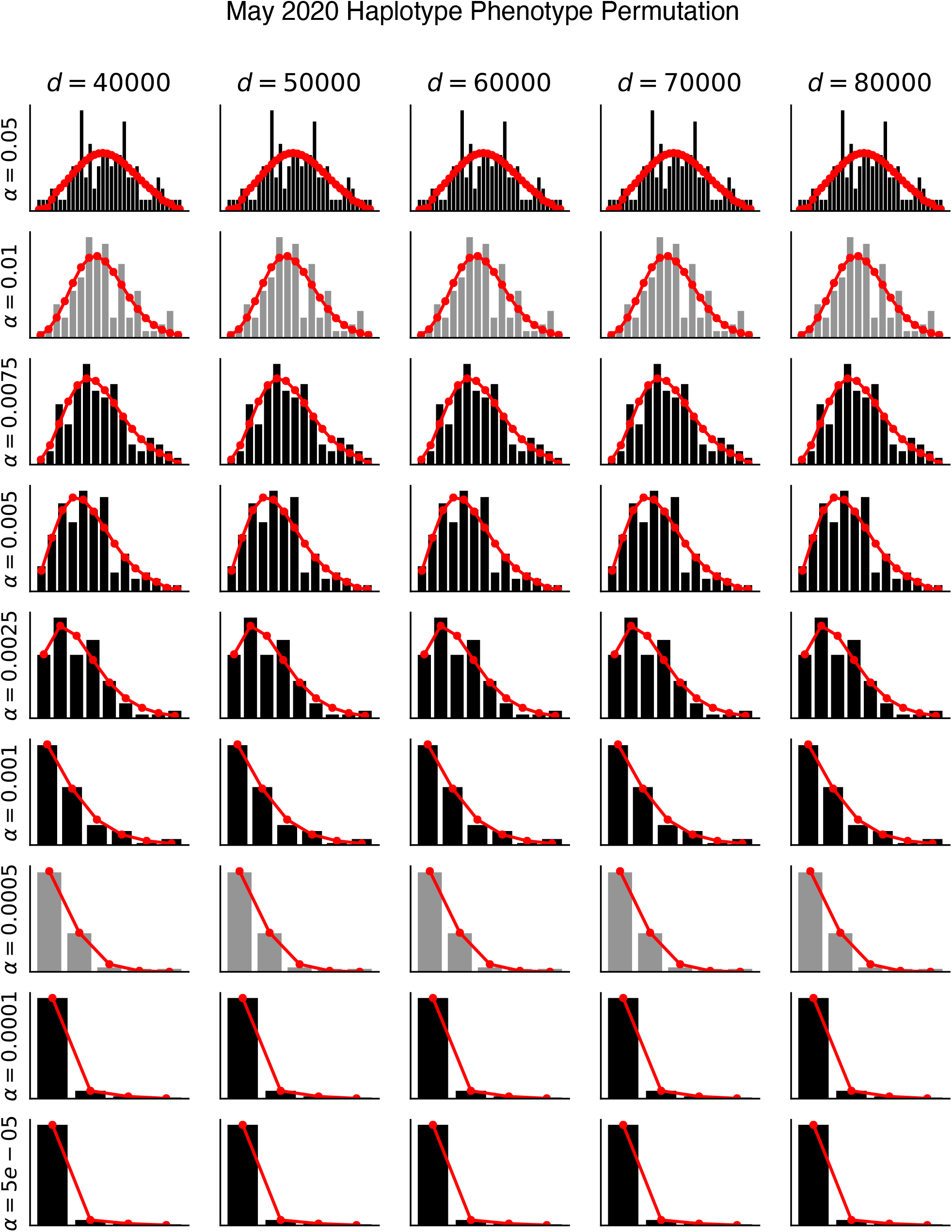
| Results of permutation testing and fits to negative binomial distributions for **(a)** April-2020 phenotype permutations, **(b)** April-2020 SNP permutations, **(c)** May-2020 phenotype permutations, **(d)** May-2020 SNP permutations, **(e)** Haplotype SNPs-only April-2020 phenotype permutations, and **(f)** Haplotype SNPs-only May-2020 phenotype permutations. Histograms indicate the number of permutations with X significant hits (black/grey bars). Negative binomial fits are shown in red (see *Methods*). Chi-squared goodness-of-fit tests were performed for each distribution. Distributions which passed the goodness-of-fit test (p > 0.05) are shown in black and those that failed (p ≤ 0.05) are shown in grey. Results are visualized for 5 distances (columns) and 9 alpha thresholds (rows). All fits are available as supplement data.

**Figure S3.**
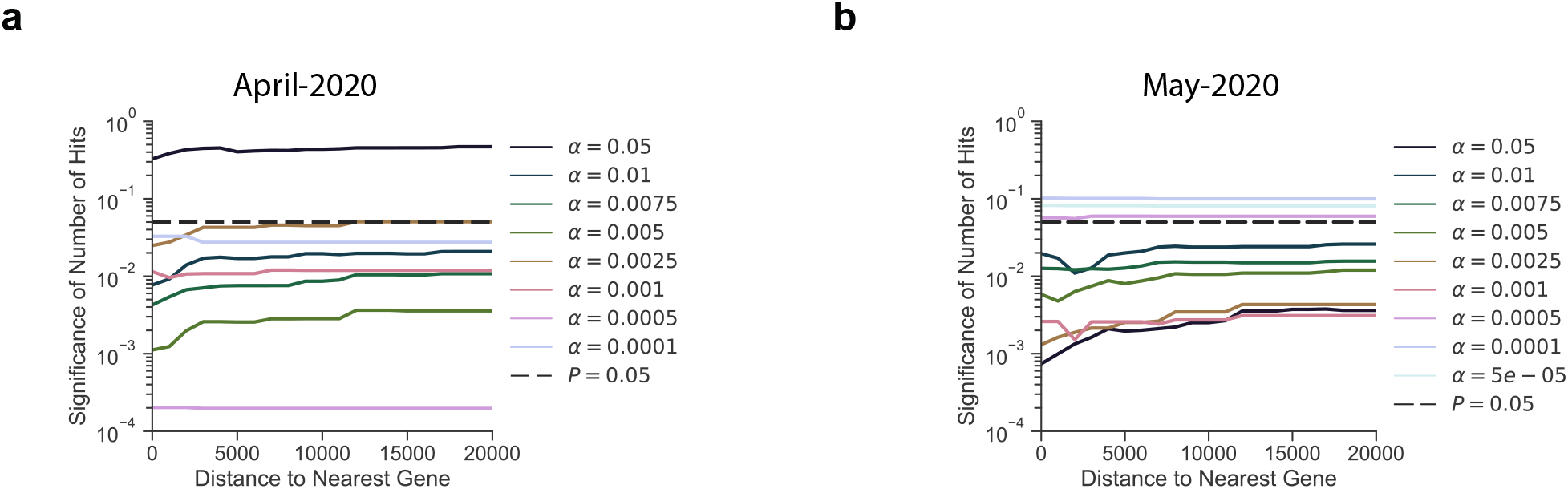
| *P*valuesfrom a Negative Binomial distribution fit to permutation of case:control phenotypes generated under the null hypothesis are shown for the Haplotype SNPs-only analyses using the April-2020 (**a**) or May-2020 (**b**) cohort. a and distance pairs as indicated; for more information see *Methods*.

**Figure S4.**
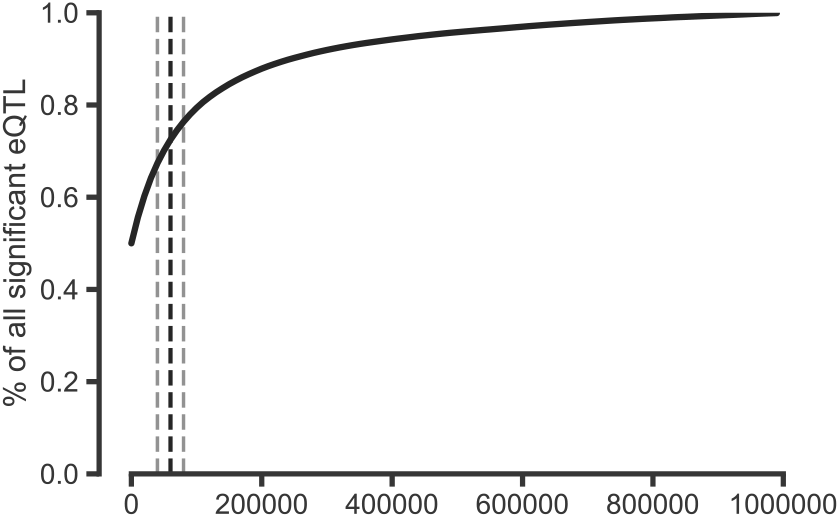
| Percent of significant eQTLs within a given distance of the gene body. Significant eQTLs were downloaded from the GTEx Portal website for Esophagus, Lung, and Heart tissues (9 tissues total) and used the provided significance thresholds to determine significance. Shown is the percent of significant eQTLs that are within X base pairs of their target gene aggregated over 9 tissues. Over 70% of significant eQTLs are within 60 Kb of their target gene. Black dashed line represents 60 Kb, grey lines represent 40 and 80 Kb.

**Figure S5.**
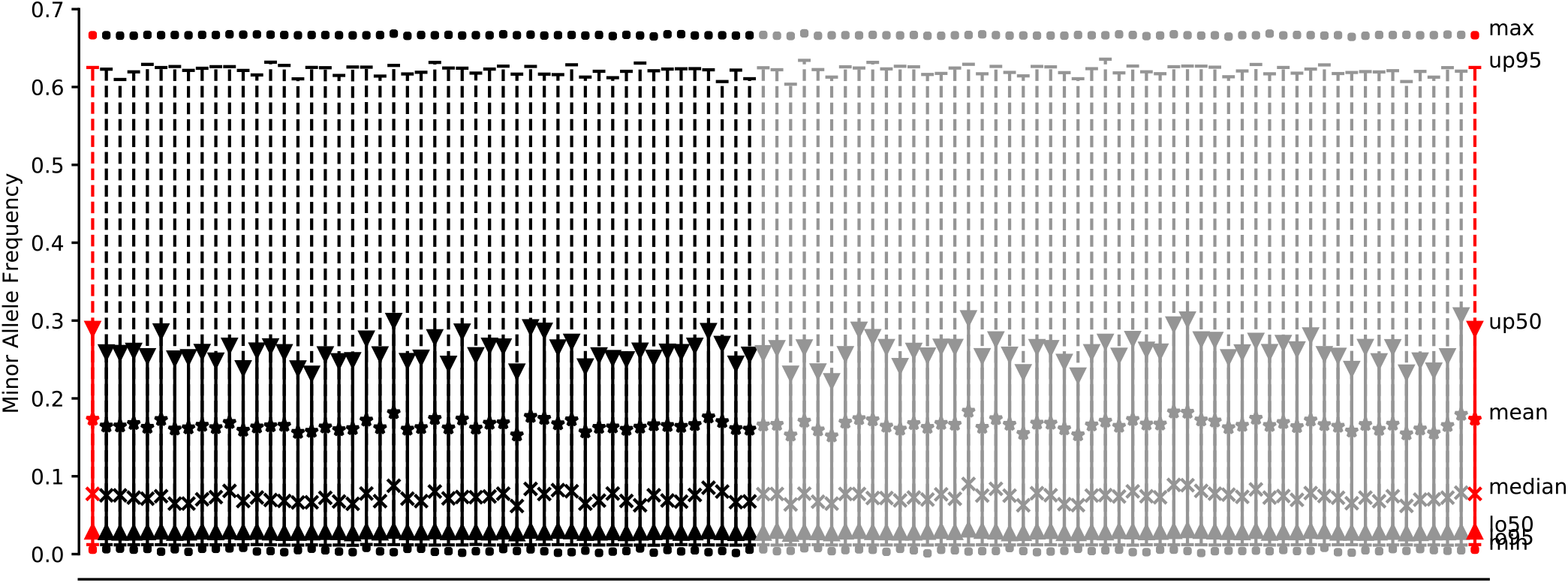
| Comparison of MAF distributions across sampled SNP sets. The medians, means, interquartile range, 95% confidence interval, minimum, and maximum are shown for each of the 100 samples of SNP sets (see *Empirical Permutation Evaluation to set Study-wide Alpha Thresholds* for details). Also shown are the same distribution statistics for the SNP set within 60Kb of complement and coagulation gene bodies (red). Each of the 100 sampled SNP sets MAF distributions were compared to the study SNP set and tested for differences using a two-sample Mann-Whitney U test. Those that were not significantly different (p > 0.05) are shown in black. Those that are significantly different (p ≤ 0.05) are shown in grey and were dropped from the analysis.

## Methods

### Ethics and Data Governance Approval

The study is approved by the Columbia University Irving Medical Center Institutional Review Board (IRB# AAAL0601) and the requirement for an informed consent was waived. A data request associated with this protocol was submitted to the Tri-Institutional Request Assessment Committee (TRAC) of New- York Presbyterian, Columbia, and Cornell and approved. The research on the UK Biobank data has been conducted using the UK Biobank Resource under Application Number 41039. The transcriptomics analysis samples were collected and processed through the Weill Cornell Medicine Institutional Review Board (IRB) Protocol 19-11021069.

#### Retrospective Clinical Study

### Cohort and Study Description

In this observational cohort study, we used a data warehouse derived from electronic health records (EHRs) from 11,116 patients treated at New York-Presbyterian/Columbia University Irving Medical Center for suspected cases of SARS-CoV-2 infection. For these patients we collected contemporary data from their current encounter (i.e. the encounter associated with their suspected SARS-CoV-2 infection) as well as historical data, if available, from their previous encounters. Contemporary data (data collected between February 1, 2020 and April 12, 2020) included insurance billing information, laboratory measurements, procedures, and SARS-CoV-2 diagnostic test results. These data were derived from the data warehouse tables in Epic. 6,927 patients have historical data (data collected prior to September 24, 2019) available from an OMOP v5 instance stored using MySQL, which included all of the standard tables for recording condition, procedure, medication, and measurement data (among others). Of these we used the insurance billing information from the condition occurrence table and demographics from the person table. See *Preparation of data for modeling* for further details on data preparation.

We used the contemporary data to define inclusion criteria and outcomes (requiring mechanical respiration and mortality) and used historical data to define patient comorbidities. We defined three hypothesized comorbidity covariates, macular degeneration, complement deficiency disorders, and disorders of coagulation. We used historical data to define these comorbidities, age, and sex. We did not include race and ethnicity data in the modeling as we have previously found issues with the data quality^28^. The race/ethnicity data we do have is included in the tables for reference. We also modeled other comorbidities previously associated with morbidity and mortality (Zhou et al and others), including history of cardiovascular disease, hypertension, obesity, and diabetes (Table 1, Table S1) -- all derived from the historical data. Coded covariate definitions, as well as lists of which diagnosis codes are most common in each group, are available in the supplemental materials and methods. We used established institutional procedures and an institutional clinical data warehouse to extract all data from the EHR.

### Defining patient outcomes

Outcome definitions were defined by data derived from the electronic health record between February 1, 2020 and April 12, 2020. Mortality is derived from a death note filed by a resident or primary provider that records the date and time of death. Intubation was used as an intermediary endpoint and is a proxy for a patient requiring mechanical respiration. We used note types that were developed for patients with SARS-CoV-2 infection to record that this procedure was completed. We validated outcome data derived from notes against the patient’s medical record using manual review.

### Preparation of data for modeling

We used MySQL and python libraries (pymysql, pandas) to extract and prepare the data for modeling. The code for data preparation is available in the github (https://github.com/tatonettilab/complementcovid) as a Jupyter Notebook titled Data Setup. We begin by creating a master list of suspected covid patients. These are patients that are either diagnosed with the disease, as indicated by a ICD10 code for SARS-CoV-2 infection, in their billing data or a patient that was tested for the presence of the virus using RT-PCR as indicated by a “lab” order for the test. We found 2,821 using the former method and 11,116 patients using the latter. We then extracted birthdates, death dates (if the patient had died or a null value otherwise), and sex codes (1 for female, 2 for male). Patients which had sex codes for non-binary genders were excluded from our analysis. We then define a “first diagnosis date” for each patient as either their first diagnosis date (by billing code) or the first date that they tested positive for SARS-CoV-2, whichever comes first. Next, we calculate each patient’s age at the time of this “first diagnosis date.” Each of the outcomes and covariates are extracted from their respective tables as detailed in the github. Whenever possible, we use the highest-level ancestor code (from the structured vocabulary in OMOP) that represents the concept we want to model. We then use the concept ancestor tables to grab all the descendant codes. Note that diabetic kidney disease was considered for inclusion and so is represented in the data preparation script, however, it was never modeled. Cough is included as a covariate as a reference symptom for comparison. The last step in the preparation process was to compute the censor dates. To do, we iterated through each patient in our master list and computed their time (in days) to intubation (if they required mechanical respiration) or death (if they died). If not, then the study end date (April 25, 2020) was used as the patient’s censored time (in days). Finally, for any patients that were not SARS-CoV-2 positive, their time-to-event values were set to a null indicator to be dropped from the dataset later. Finally, the data are all combined in a pandas (version 1.0.3) dataframe and saved to disk as a pickle file for efficient loading.

### Statistical Model

Our patient timelines may be censored since our study cohort included patients that were being treated at the time of analysis. We performed survival analysis on the intubation orders and death using a Cox proportional-hazards model and visualized the risk using Kaplan-Meier curves using the lifelines python package (version 0.24.4). Error estimates on the Kaplan-Meier curves are estimated using Greenwood’s Exponential Formula^29^. We fit both univariate models and models fit on the covariate, age, and sex and used log-likelihood to assess significance. We reported Cox proportional hazards coefficients and their 95% confidence intervals (Table 1). We modeled whether or not a patient had macular degeneration, a complement deficiency disorder, or a coagulation disorder as binary variables (1=yes, 0=no). Code definitions provided in Table S1. We also included other significant comorbidities suggested by previous studies, CAD, hypertension, T2DM, obesity, or smoking status as binary variables (1=yes, 0=no), sex as a binary variable (0=female, 1=male), age as quantitative variable, older age over 65 (note that age over 65 is used *only* for illustrative purposes and is not used in multivariate modeling -- in the multivariate model age as a quantitative variable is used), and outcome as a binary variable (1=yes, 0=no). The outcome of interest was coded as 0 until the day it occurred (the date of the first intubation order following admission or the death date) or the date of analysis, whichever occurred first. Survival curves are generated for the indicated variables by setting all other variables to their respected averages within the training data. Note that we dropped patients who experienced the outcome before their initial diagnosis. This is either due to patients being hospitalized prior to infection (in the case of intubation) or errors in the coded data. We dropped 121 patients for intubation prior to infection and 12 patients for prior death. We also restricted the study to 90 days from the start date. One patient was removed for having an event outside of this range.

### Covariate Correlations

Using the data prepared as discussed above, we computed pairwise statistical correlations between age, sex as well as history of macular degeneration, complement deficiency disorders, coagulation disorders, HTN, T2DM, obesity, and CAD. We computed them using data from all suspected patients (tested both positive and negative) as well as only those patients who tested positive. We used spearman rho and the tanimoto coefficients (1-Jaccard distance) as our measures of correlation. For the comparison using the tanimoto coefficient we binarized age as greater than or equal to 65.

### Statistical Software

We used Jupyter Notebooks (jupyter-client version 5.3.4 and jupyter-core version 4.6.1) running Python 3.7 and all fit models using the python lifelines package (version 0.24.4).

#### Transcriptomic Analysis of NP swabs

### Sample Collection and Processing

Patient specimens were collected with patients’ consent at New York Presbyterian Hospital (NYPH) and then processed for RT-PCR as described previously^30^. Nasopharyngeal (NP) swab specimens were collected using the BD Universal Viral Transport Media system (Becton, Dickinson and Company, Franklin Lakes, NJ) from symptomatic patients.

### Extraction of Viral RNA and RT-PCR detection

Total viral RNA was extracted from deactivated samples using automated nucleic acid extraction on the QIAsymphony and the DSP Virus/Pathogen Mini Kit (QIAGEN). One step reverse transcription to cDNA and real-time PCR (RT-PCR) amplification of viral targets, E (envelope) and S (spike) genes and internal control, was performed using the Rotor-Gene Q thermocyler (QIAGEN).

### Human Transcriptome Analysis

RNA-seq reads that mapped unambiguously to the human reference genome via Kraken2 were used to detect transcriptional responses to SARS-CoV-2 infection as described previously^30^. Briefly, reads were trimmed with TrimGalore, aligned with STAR (v2.6.1d) to the human reference build GRCh38 and the GENCODE v33 transcriptome reference, gene expression was quantified using featureCounts, stringTie and salmon using the nf-core RNAseq pipeline. Sample QC was reported using fastqc, RSeQC, qualimap, dupradar, Preseq and MultiQC. Reads, as reported by featureCounts, were normalized using variance-stabilizing transform (vst) in DESeq2 package in R and DESeq2 was used to call differential expression with either Positive cases vs Negative, or viral load (High/Medium/Low/None) as reported by RT-PCR cycle threshold (Ct) values. Transcript counts (per million) were used to rank genes and perform gene set enrichment analysis (GSEA).

### Reverse Transcriptase, quantitative real-time PCR (RT-PCR)

The presence of SARS-CoV-2 in clinical samples was determined by RT-PCR. Briefly, primers for the E (envelope) gene (which detects all members of the lineage B of beta-CoVs), and the S (spike) gene (which specifically detect SARS-CoV-2). Samples were annotated using RT-PCR cycle threshold (Ct) value for SARS-CoV-2 primers as follows: Ct ≤ 18 were assigned “high viral load"; Ct 18 - 24 were assigned “medium viral load”; and Ct 24 - 40 were assigned “low viral load” stratifications; Ct > 40 was classified as negative (-).

#### Genetic Analysis of UK Biobank

### Data Source

UK Biobank subjects that were of White British descent, in the UK Biobank PCA calculations and therefore without 3rd degree and above relatedness and without aneuploidy, were used in this study, totaling 337,147 subjects (181,032 females and 156,115 males) (Bycroft 2018). Of the nearly 500,000 participants, approximately 50,000 subjects were genotyped on the UK BiLEVE Array by Affymetrix while the rest were genotyped using the Applied Biosystems UK Biobank Axiom Array, with over 800,000 markers using build GRCh37 (hg19). The arrays share 95% marker coverage. We extracted markers with a minor allele frequency greater than 0.005, INFO score greater than 0.3, and Hardy-Weinberg equilibrium test mid-p value greater than 10-10 using PLINK2^31^. UKBB version 3 Imputation combined the Haplotype Research Consortium with the UK10K haplotype resource using the software IMPUTE4 (UK Biobank White paper). Association analyses were performed using a logistic regression model with additive gene dosage and covariates including age at 2018, sex, first 10 principal components (provided by the UK Biobank), and the genotyping array the sample was carried out on. We determined the alpha threshold for study-wide significance using an empirical permutation analysis (see *Empirical Permutation Evaluation to set Alpha Thresholds*). We performed a study-wide association analysis comparing variants for subjects that were SARS-CoV-2 positive and required hospitalization against the entire population of 337,147 subjects

### Targeted Gene Set Definition

The union of coagulation and complement related gene sets (with immunoglobulin genes removed) that are part of MsigDB was used to define the set of 102 genes used in this study. For each gene, we used the transcriptional start and stop site from the hg19 build of the human genome to define a catchment window of 80kbp. From the 805,426 variants profiled in the UK Biobank genotyping data after quality control and QC filters using PLINK2 (see above), 3,540 variants within the transcribed region of the genes of interest or within 80kbp flanking the transcribed region, 2,888 are within 60kbp, 2,292 are within 40kbp, and 936 are located in haplotype blocks with study genes.

### Empirical Permutation Evaluation to set Study-wide Alpha Thresholds

We used permutation to estimate null distributions of the number of hits expected at 9 alpha thresholds varying from (5×10^-5^ to 0.05) and by varying the distance threshold from 40kb to 80kb. As shown previously, 80% of GWAS hits are within 60Kb of the nearest gene^32^. Further, as shown in Supplemental Figure S4, we empirically determined that the majority of eQTLs (>70%) are within 60kb of gene bodies. We performed two sets of permutation analyses: (i) permuted the initial set of genes on which the included variant loci were chosen and (ii) permuted the case/control labels. We repeated each 100 times and used the resulting data to fit a negative binomial distribution as our estimate of the null. Additionally, we evaluated each of the sampled SNP variant sets from (i) and compared their MAF distribution with the MAF distribution of the Complement and Coagulation set. We removed any sets that were significantly different (nominal p-value < 0.05) according to a Mann-Whitney U test (52 of 100 sets were removed due to this criterion; see Supplemental Figure S5). We found that the negative binomial fit the data the best according to a goodness of fit test (Supplemental Figure S2). We used this distribution to assess statistical significance for each combination of alpha and distance values. The result is two estimates of the significance for each alpha (α), distance (d) pair, 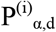 and 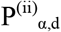, from permutation analyses (i) and (ii) above, respectively. For example:

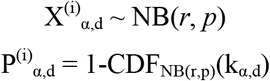

where 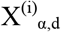 is the number of permutation loci with a p-value under the threshold, α. The parameters *r* and *p* of the negative binomial represent the number of successes/failures and the probability of success, respectively. Both *r* and *p* are fit using non-linear least squares (the curve_fit function in scipy.optimize) on 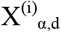, the count data from the permutation analyses for the given α and d. The P is then calculated using the CDF of the fitted negative binomial distribution.

For the gene set permutation analysis (i.e. (i) above) we evaluated each of the 100 replicates to confirm that the minor allele frequency distribution was statistically indistinguishable from that of the complement and coagulation gene set variants. We did so by performing a Mann-Whitney U test between the two distributions and excluded any replicates that showed a significant difference (nominal p-value < 0.05). 52 replicates were excluded because of this requirement (Figure SX). This MAF distribution analysis is not necessary for the case/control permutation analysis (i.e. (ii) above) as the loci are the same in each replicate and it is the case/control labels that are permuted.

Finally, to set the study-wide alpha for each study we chose the greatest threshold value that was gave a P of 0.05 or less for both permutation analysis method:

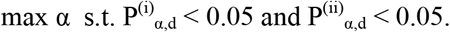

Finally, this entire process was repeated for two cohorts of patients, (a) the initial COVID cohort released by the UK Biobank in April 2020 and (b) the updated COVID cohort released in May 2020. The chosen a for April was 0.001 and the chosen α for May was 0.0025. A data file of all of the distribution fit results and their resulting chi-squared goodness-of-fit statistics is made available in the supplemental materials.

We also performed this permutation significance estimation for the haplotype-derived SNP sets although the distances for all loci chosen using that method are below the minimum in this analysis of 40Kb so those results are constant with regards to distance (Figures S3a-b). The chosen α for the LD-derived SNP sets is 0.01 and 0.0075 for April and May, respectively.

### Haplotype block-based selection of SNPs

We identified haplotype blocks based on linkage disequilibrium within the UK Biobank data genotype data of the 337,147 subjects using PLINK1.9, where the lower 90% confidence interval is greater than 0.70 and the upper 90% confidence interval is at least 0.98. We identified blocks of interests, and subsequently the variants within those blocks, as those that contain any part of the genes of interest as denoted by the transcriptional start and end sites from the hg19 build of the human genome. From the 805,426 variants profiles in the UK Biobank genotype data, we identified 7,281 variants within the genes of interest. After applying additional QC filters using PLINK2, 936 variants remained for analysis.

### Software

We used PLINK v2.00a2LM 64-bit Intel (26 Aug 2019) to run the genetic association analysis. We used PLINK v1.90b6.10 64-bit (17 Jun 2019) to identify haplotype blocks based on linkage disequilibrium. We used Jupyter Notebooks (jupyter-client version 5.3.4 and jupyter-core version 4.6.1) running Python 3.7, numpy 1.18.1, and scipy 1.4.1 for the permutation analyses.

## References

1 Zhang, L. et al. Crystal structure of SARS-CoV-2 main protease provides a basis for design of improved alpha-ketoamide inhibitors. Science 368, 409–412, doi:10.1126/science.abb3405 (2020).

2 Dai, W. et al. Structure-based design of antiviral drug candidates targeting the SARS-CoV-2 main protease. Science, doi:10.1126/science.abb4489 (2020).

3 Gordon, D. E. et al. A SARS-CoV-2-Human Protein-Protein Interaction Map Reveals Drug Targets and Potential Drug-Repurposing. bioRxiv, 2020.2003.2022.002386, doi:10.1101/2020.03.22.002386 (2020).

4 Chen, G. et al. Clinical and immunological features of severe and moderate coronavirus disease 2019. J Clin Invest, doi:10.1172/JCI137244 (2020).

5 Moore, B. J. B. & June, C. H. Cytokine release syndrome in severe COVID-19. Science, doi:10.1126/science.abb8925 (2020).

6 Lasso, G. et al. A Structure-Informed Atlas of Human-Virus Interactions. Cell 178, 1526-1541 e1516, doi:10.1016/j.cell.2019.08.005 (2019).

7 Merle, N. S., Church, S. E., Fremeaux-Bacchi, V. & Roumenina, L. T. Complement System Part I - Molecular Mechanisms of Activation and Regulation. Front Immunol 6, 262, doi:10.3389/fimmu.2015.00262 (2015).

8 Holers, V. M. Complement and its receptors: new insights into human disease. Annu Rev Immunol 32, 433–459, doi:10.1146/annurev-immunol-032713-120154 (2014).

9 Liszewski, M. K., Java, A., Schramm, E. C. & Atkinson, J. P. Complement Dysregulation and Disease: Insights from Contemporary Genetics. Annu Rev Pathol 12, 25–52, doi:10.1146/annurev-pathol-012615-044145 (2017).

10 Wu, J. & Sun, X. Complement system and age-related macular degeneration: drugs and challenges. Drug Des Devel Ther 13, 2413–2425, doi:10.2147/DDDT.S206355 (2019).

11 Ambati, J., Atkinson, J. P. & Gelfand, B. D. Immunology of age-related macular degeneration. Nat Rev Immunol 13, 438–451, doi:10.1038/nri3459 (2013).

12 Degn, S. E., Jensenius, J. C. & Thiel, S. Disease-causing mutations in genes of the complement system. Am J Hum Genet 88, 689–705, doi:10.1016/j.ajhg.2011.05.011 (2011).

13 Zhou, F. et al. Clinical course and risk factors for mortality of adult inpatients with COVID-19 in Wuhan, China: a retrospective cohort study. Lancet 395, 1054–1062, doi:10.1016/S0140-6736(20)30566-3 (2020).

14 Nicholson-Weller, A. & Wang, C. E. Structure and function of decay accelerating factor CD55. J Lab Clin Med 123, 485-491 (1994).

15 Bycroft, C. et al. The UK Biobank resource with deep phenotyping and genomic data. Nature 562, 203–209, doi:10.1038/s41586-018-0579-z (2018).

16 Sudlow, C. et al. UK biobank: an open access resource for identifying the causes of a wide range of complex diseases of middle and old age. PLoS Med 12, e1001779, doi:10.1371/journal.pmed.1001779 (2015).

17 Smith, N. L. et al. Genetic predictors of fibrin D-dimer levels in healthy adults. Circulation 123, 1864–1872, doi:10.1161/CIRCULATIONAHA.110.009480 (2011).

18 Han, X. et al. Genome-wide meta-analysis identifies novel loci associated with age- related macular degeneration. J Hum Genet, doi:10.1038/s10038-020-0750-x (2020).

19 Consortium, G. T. et al. Genetic effects on gene expression across human tissues. Nature 550, 204–213, doi:10.1038/nature24277 (2017).

20 Rehman, A. A., Ahsan, H. & Khan, F. H. alpha-2-Macroglobulin: a physiological guardian. J Cell Physiol 228, 1665–1675, doi:10.1002/jcp.24266 (2013).

21 Gary-Bobo, M., Nirde, P., Jeanjean, A., Morere, A. & Garcia, M. Mannose 6-phosphate receptor targeting and its applications in human diseases. Curr Med Chem 14, 29452953, doi:10.2174/092986707782794005 (2007).

22 Ermert, D. & Blom, A. M. C4b-binding protein: The good, the bad and the deadly. Novel functions of an old friend. Immunol Lett 169, 82–92, doi:10.1016/j.imlet.2015.11.014 (2016).

23 Goeijenbier, M. et al. Review: Viral infections and mechanisms of thrombosis and bleeding. J Med Virol84, 1680-1696, doi:10.1002/jmv.23354 (2012).

24 Nascimento, E. J. et al. Alternative complement pathway deregulation is correlated with dengue severity. PLoS One 4, e6782, doi:10.1371/journal.pone.0006782 (2009).

25 Pastor, A. F. et al. Complement factor H gene (CFH) polymorphisms C-257T, G257A and haplotypes are associated with protection against severe dengue phenotype, possible related with high CFH expression. Hum Immunol 74, 1225–1230, doi:10.1016/j.humimm.2013.05.005 (2013).

26 Risitano, A. M. et al. Complement as a target in COVID-19? Nat Rev Immunol, doi:10.1038/s41577-020-0320-7 (2020).

27 Mastaglio, S. et al. The first case of COVID-19 treated with the complement C3 inhibitor AMY-101. Clin Immunol 215, 108450, doi:10.1016/j.clim.2020.108450 (2020).

28 Polubriaginof, F. C. G. et al. Challenges with quality of race and ethnicity data in observational databases. J Am Med Inform Assoc 26, 730–736, doi:10.1093/jamia/ocz113 (2019).

29 Hosmer, D. W., Lemeshow, S. & May, S. Applied survival analysis: regression modeling of time-to-event data. 2nd edn, (Wiley-Interscience, 2008).

30 Butler, D. J. et al. Shotgun Transcriptome and Isothermal Profiling of SARS-CoV-2 Infection Reveals Unique Host Responses, Viral Diversification, and Drug Interactions. bioRxiv, 2020.2004.2020.048066, doi:10.1101/2020.04.20.048066 (2020).

31 Chang, C. C. et al. Second-generation PLINK: rising to the challenge of larger and richer datasets. Gigascience 4, 7, doi:10.1186/s13742-015-0047-8 (2015).

32 Brodie, A., Azaria, J. R. & Ofran, Y. How far from the SNP may the causative genes be? Nucleic Acids Res 44, 6046–6054, doi:10.1093/nar/gkw500 (2016).

